# Detection of Atrial Fibrillation with a Hybrid Deep Learning Model and Time-Frequency Representations

**DOI:** 10.1101/2025.06.03.25328929

**Authors:** Yu Luo, Baixiang Huang, Jingzi Zhu, Xueying Zeng, Qing Zhang

## Abstract

Atrial fibrillation (AF), a common cardiac arrhythmia, can lead to severe complications, emphasizing the urgent need for effective detection methods. This study proposes an automated algorithm for AF detection that combines time-frequency analysis with deep learning techniques, achieving exceptional performance across multiple public ECG datasets. The proposed system applies variational mode decomposition (VMD) to decompose non-stationary ECG signals, followed by Hilbert transform (HT) to generate time-frequency representations. These 2D maps are then processed by a deep learning model for classification. We introduce a novel architecture, SwinMobileNet, which integrates the strengths of the Swin Transformer and MobileNetV2 to effectively model both spatial and temporal features in ECG signals. An adaptive attention mechanism ensures efficient and accurate classification. The implementation of this algorithm is available at: https://anonymous.4open.science/r/SwinMobileNet1-0C3C.

## I. INTRODUCTION

**A** Trial fibrillation (AF), a common persistent arrhythmia, is a global concern. In 2019, there were around 59.5 million AF patients globally, with incidence rising with age [1]. Low-income countries face challenges due to limited resources, aging populations, and prevalent risk factors like hypertension and obesity. Treatment options include drugs, device therapy, and surgery. The global pulsed field ablation market size was estimated at USD 913.1 million in 2024 and is projected to grow at a CAGR of 33.1% from 2025 to 2030. [2]

AF can be detected by multiple methods like ECG and Holter. A conventional ECG records cardiac electrical activity changes for a few to ten seconds. Holter recordings last from 14 to 72 hours. ECG reveals AF features such as P wave loss and F wave appearance, yet has drawbacks like interference susceptibility and inability to comprehensively assess cardiac function [3]. For paroxysmal AF, traditional methods with short recording times often miss patients with infrequent episodes.

Research has demonstrated the effectiveness of convolutional neural networks (CNNs) [4], recurrent neural networks (RNNs) [5], long short-term memory networks (LSTMs) [6] in detecting atrial fibrillation. These methods can automatically analyze ECG data and improve diagnosis accuracy and efficiency. Nonetheless, they still face challenges in terms of data quality, model sensitivity to complications, and accuracy, which require further research and improvements to optimize their performance.

We consider using a time-frequency graph in conjunction with deep learning to solve these problems. We use time-frequency analysis—a signal processing method—to analyze the characteristics of signals in two dimensions: time and frequency. It overcomes the limitations of traditional signal analysis methods, which can only provide the overall frequency of the signal but cannot reflect the frequency change over time [7]. It is also convenient for multi-signal comparison and classification. Widely used and of significant value, it opens up a new research direction. Converting ECG signals into time-frequency maps allows for a more comprehensive capture of signal characteristics, enhancing diagnosis accuracy and providing rich input data for subsequent deep learning models.

In summary, this paper proposes an algorithm that combines time-frequency analysis and deep learning to solve many problems encountered in AF detection. The proposed method makes full use of the time domain and frequency domain characteristics of ECG signals. Combining variational mode decomposition (VMD) and Hilbert spectrum analysis (HT), we turned the one-dimensional ECG signal into a two-dimensional time-frequency map. This improved the noise filtering function and decreased interference from noise on the results. We tried VGG16 [8], ResNet34 [9], MobileNetV2 [10], InceptionV2 [11], Swin Transformer [12], and other models during the exploration process. We carefully analyzed and evaluated each model’s performance and characteristics, taking into account various factors, such as ECG signal processing requirements and AF detection accuracy improvements. Finally, we built a deep learning network framework using SwinTransformer and MobileNet V2. This framework automatically sorts normal time-frequency maps and AF time-frequency maps with high accuracy, which makes AF detection more accurate. The flow diagram of the proposed method is shown in Fig. 1

**Fig. 1.**
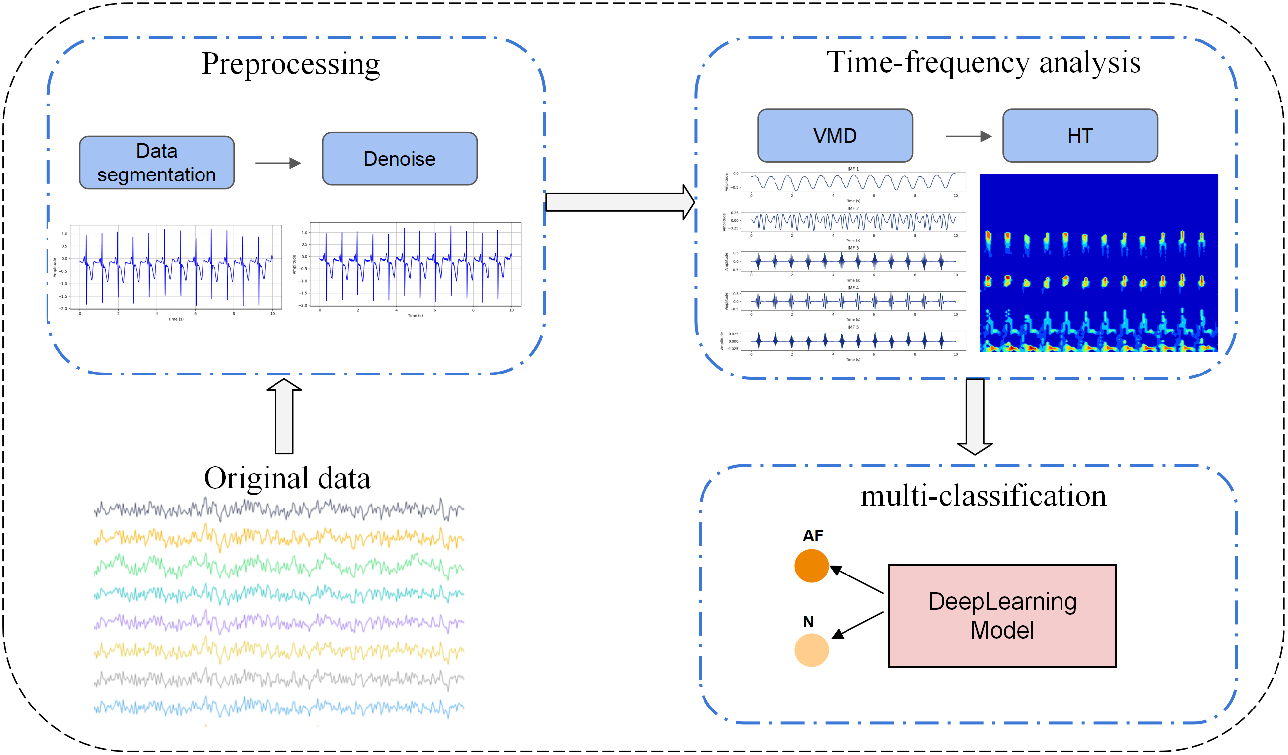
Overall framework.

**Fig. 2.**
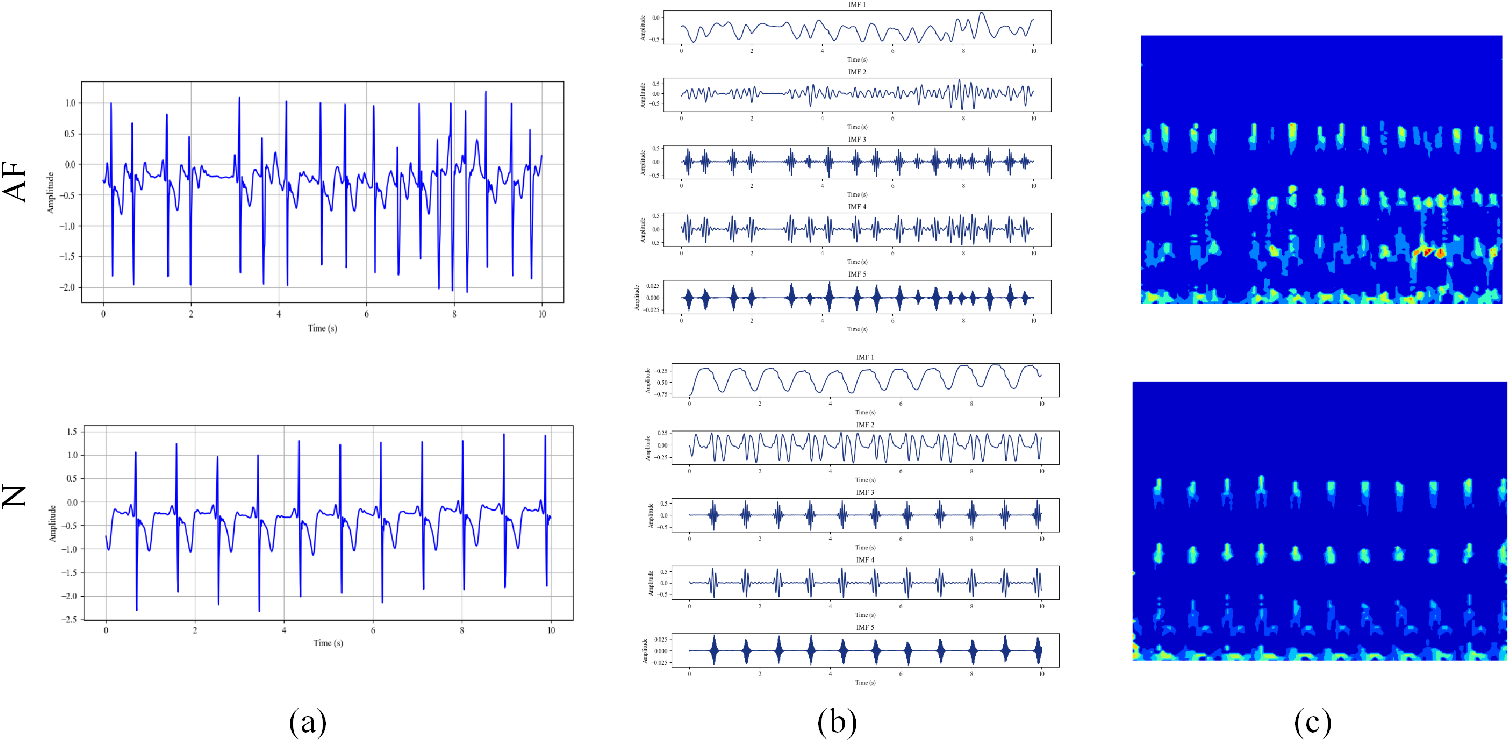
Schematic diagram of electrocardiogram conversion time-frequency diagram.

## II. RELATED WORK

### A. Existing Techniques of Deep learning Model in AF Detection

The application of deep learning technology in AF detection has gradually evolved from initial exploration to mature application. For example, by treating ECG signals as one-dimensional (1D) data, 1D-CNN is used for fast and accurate classification of electrocardiograms [13] [14] [15], but the robustness of this algorithm remains an important issue [16]. In addition, there are studies that apply an improved U-net architecture for diagnosing arrhythmias [17]. Besides 1D-CNN models, there are also models based on Recurrent Neural Networks (RNN), such as the feature-induced LSTM model used to classify five different types of arrhythmias [18]. Furthermore, a CNN and LSTM-based deep neural network model has been proposed, which classifies heartbeats using both topic- and category-oriented methods [19].

### B. Application of Deep Learning and Time-frequency Analysis

In recent years, the combination of deep learning and time-frequency analysis has introduced new ideas for AF detection. For example, in [20], the instantaneous frequency and spectral entropy of ECG signals are used as inputs for the LSTM model to classify the ECG signals. In [21] and [22], one-dimensional ECG signals are converted into two-dimensional spectral images using Short-time Fourier transform (STFT), followed by the use of 2D CNN or Bi-LSTM networks for arrhythmia classification. Additionally, in [23], the Superlet transform (SLT) is employed to convert 1D ECG signals into 2D time-frequency maps, which are then input into the DenseNet-201 architecture for multi-level arrhythmia classification. [24] explores various time-frequency analysis techniques such as STFT, Chirplet transform, Stockwell transform, and Poincaré maps, which are applied to one-dimensional pre-processed ECG recordings to extract two-dimensional patterns for atrial fibrillation recognition. Furthermore, in [25], a Hilbert-Huang Transform (HHT) is used to convert one-dimensional ECG signals into time-frequency signals, which are then processed using the DenseNet network for AF detection.

## III. METHOD

### A. Preprocessing

For ECG signals, we use wavelet denoising methods to remove high-frequency noise. First, we apply the discrete Wavelet transform (DWT) to the ECG signal, decomposing it into wavelet coefficients corresponding to different levels.

Then, we use the hard thresholding method to process the coefficients of the high-frequency subbands. The hard thresholding method sets coefficients below a certain threshold to zero, thereby suppressing high-frequency noise while preserving important signal features. The thresholding function is given by

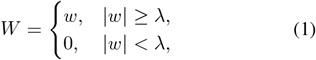

with the threshold *λ* being given by

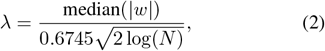

where *w* is the original wavelet coefficient, *W* is the processed wavelet coefficient, and *N* is the signal length.

Finally, the denoised coefficients are reconstructed into the denoised ECG signal through the inverse wavelet transform. Subsequently, the study focuses on two short time windows, 30 seconds and 10 seconds, of the ECG segments. That is, the filtered ECG signal is segmented, and segments of 10 seconds or 30 seconds in length are cut. The denoised ECG segments are shown in Fig. 4(a).

### B. Time-frequency analysis algorithm of ECG data

We utilize VMD to decompose the ECG signal into several IMF(*t*) components. Next, we calculate the instantaneous frequency and amplitude of each mode using the HT Finally, by combining all modal information, we obtain the time-frequency representation of the ECG signal.

#### 1) Variational Mode Decomposition

Variational mode decomposition is a self-adaptive, completely non-recursive method for modal variation and signal processing [26]. This method solves the end effect and the repetition of modal components in the EMD method [27], has more robust mathematical theoretical support, can reduce the instability of high complexity and high nonlinearity time series data, decompose it, and obtain a relatively stable sub-sequence [28]. The core idea of VMD is to construct and solve the variational problem

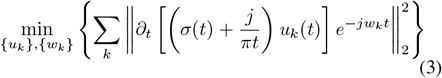

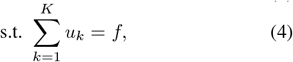

where *K* is the number of modes to be decomposed (a positive integer), {*u*_*k*_} = {*u*_1_, …, *u*_*K*_} are the *K* mode components obtained from the decomposition, {*w*_*k*_} = {*w*_1_, …, *w*_*K*_} are the frequency centers of each component, and *σ*(*·*) is the Dirac delta function.

To find the optimal solution of the constrained variational problem in equations (3) and (4), we introduce the augmented Lagrange function, i.e., equation (5),

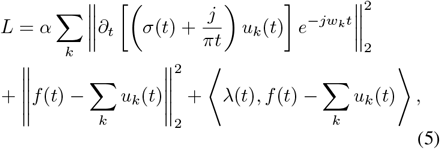

where *α* is the penalty parameter, and *λ* is the Lagrange multiplier. Solving equation (5) is equivalent to solving equation (3) and (4), thereby decomposing the original signal into *K* mode components.

The VMD decomposition process is the solution process of the variational problem. The basic steps for signal decomposition are as described in Algorithm 1.

##### Algorithm 1

Algorithm for Decomposition

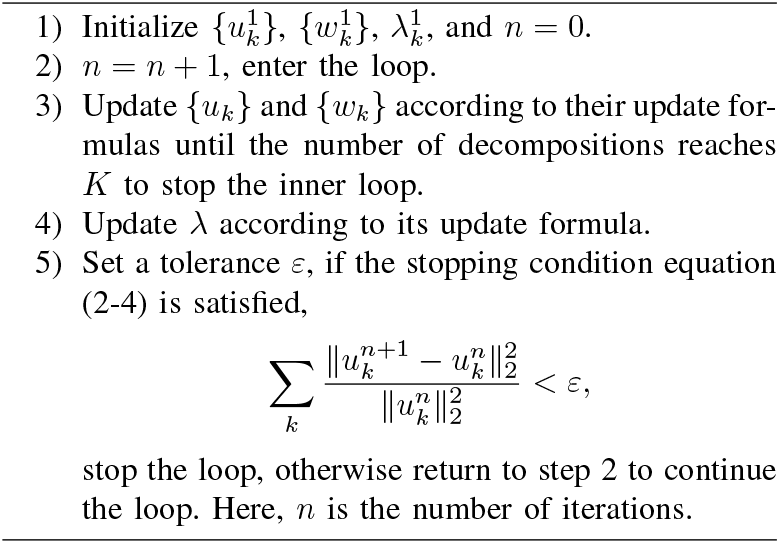

ECG signal analysis using VMD requires determining the appropriate decomposition layers. Insufficient decomposition may miss subtle information, while improper decomposition can lead to over-decomposition. Cardiac activity primarily lies in the 0.05–100 Hz range, with 80% of energy concentrated between 0.5–45 Hz. Decomposing the signal into five major modal components allows for a more precise characterization of the QRS complex. Thus, a K value of 5 is optimal for ECG signal decomposition.The modal components obtained after VMD decomposition are shown in Fig. 4(b).

#### 2) Hilbert-Huang transformation

The ECG signal was decomposed by VMD to obtain five modal components.

Hilbert transform is applied to the intrinsic mode function as follows:

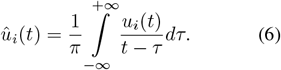

The instantaneous frequency formula is obtained as follows:

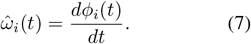

The instantaneous amplitude formula is obtained as follows:

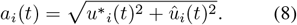

The original signal is transformed into the form of instantaneous frequency and instantaneous amplitude as follows:

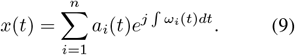

Combining the instantaneous frequencies and amplitudes of all IMFs, a 2-dimensional time-frequency graph can be obtained. The resulting time-frequency diagram is shown in Fig. 4(c).

### C. Time frequency graph classification model

In this section, we first present the overall structure of the time-frequency graph classification model based on SwinTransfomer [29] versus MobileNet [30]. Subsequently, we elaborate on two parallel network architectures. Finally, we will describe the loss function used during training.

#### 1) Overall Architecture

In image classification tasks, convolutional neural networks (CNNs), such as MobileNetV2, have demonstrated considerable efficiency and computational advantages, particularly in resource-constrained environments. However, CNNs face inherent limitations due to their restricted receptive field, which makes it challenging to capture long-range dependencies within the data. On the other hand, Transformer-based architectures, like the Swin Transformer, excel in modeling global context by leveraging self-attention mechanisms, enabling them to capture long-range dependencies more effectively.

Motivated by the complementary strengths of these two approaches, we propose a novel parallel integrated architecture, SwinMobileNet (as shown in Figure 3), which combines the power of the Swin Transformer and MobileNet. This hybrid design aims to strike an optimal balance between computational efficiency and feature expressiveness. By incorporating both local and global feature extraction mechanisms, it allows for more effective fusion of local and global information, ultimately enhancing the model’s capability in image classification tasks. Additionally, we introduce a custom spatial-channel attention mechanism that adaptively emphasizes critical spatial regions and feature channels, further improving the model’s ability to capture relevant information.The architecture specifications of SwinMobileNet are detailed in Table I.

**Table I.** SwinMobileNet Architecture Specifications

| Layer name | Input size (C,H,W) | Settings | Output size (C,H,W) |
| --- | --- | --- | --- |
| MobileNetV2 Branch |  |  |  |
| Input | (3, 224, 224) | Image input | (3, 224, 224) |
| Features Part1 | (3, 224, 224) | MobileNetV2 features (without last layer) | (320, 7, 7) |
|  | (320, 7, 7) | SE-Attention + Custom Attention | (320, 7, 7) |
| Features Part2 | (320, 7, 7) | Last MobileNetV2 conv layer | (1280, 7, 7) |
| Swin Transformer Branch |  |  |  |
| Swin Block | (3, 224, 224) | Patch Embedding (patch size=4) | (96, 56, 56) |
| | | Swin Transformer Block × 2 | (192, 28, 28) |
| | | Swin Transformer Block × 6 | (384, 14, 14) |
| | | Swin Transformer Block × 2 | (1280, 7, 7) |
| Feature Fusion and Classification |  |  |  |
| Fusion | (1280, 7, 7)<br>(1280, 7, 7) | Element-wise addition | (1280, 7, 7) |
| Global Pool | (1280, 7, 7) | Global average pooling | (1280, 1, 1) |
| Classifier | (1280) | Dropout(0.2) | (1280) |
|  | (1280) | Linear | (3) |

**Fig. 3.**
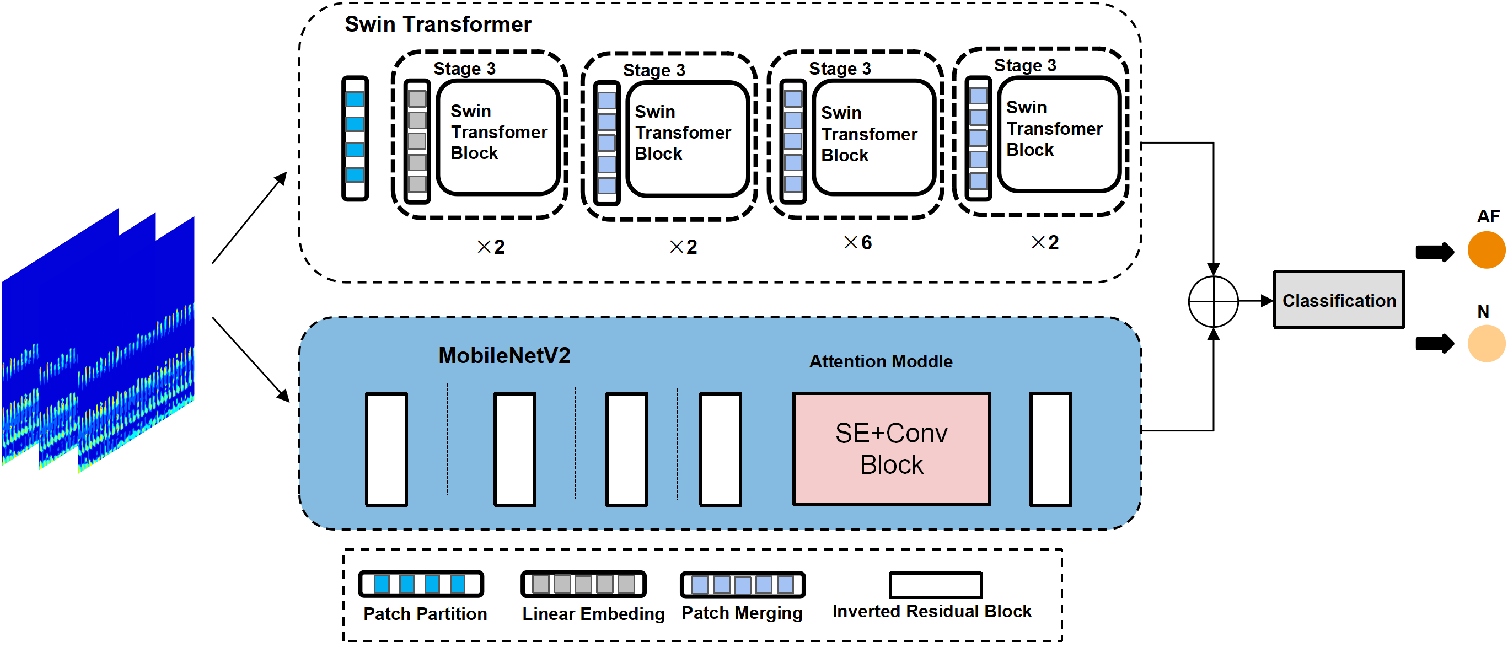
The Network Framework of SwimMobileNet.

**Fig. 4.**
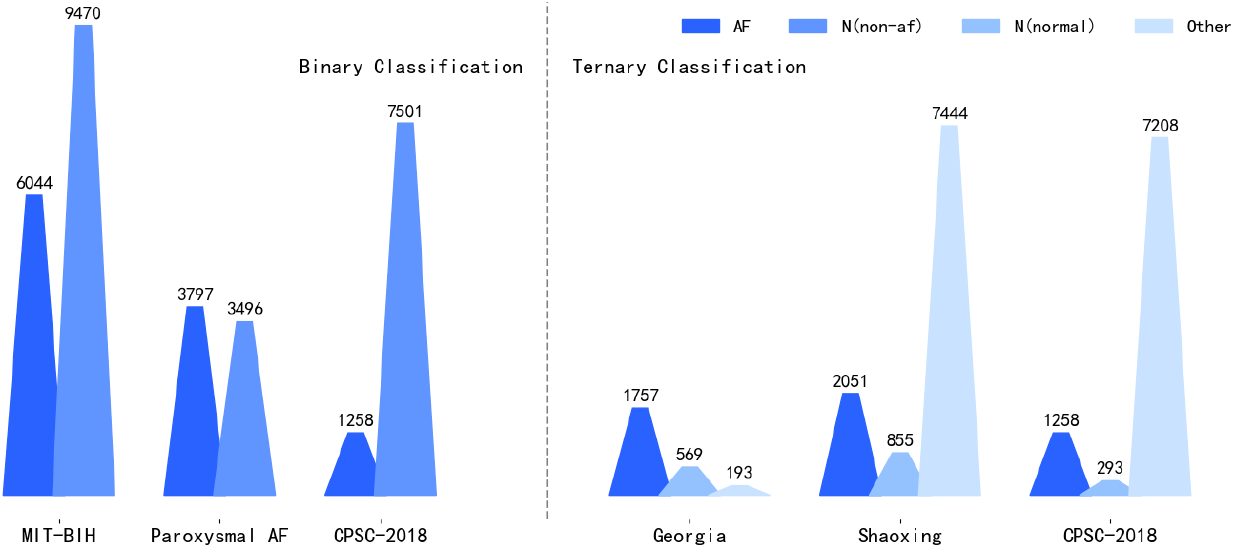
Dataset Distribution.

#### 2) MobileNet and Swin Transformer for Extracting Multilevel Features

##### MobileNet Part

We adopt the MobileNetV2 architecture, whose weights are pretrained on ImageNet, and divide it into two stages: the early feature extraction module and the later feature extraction module, with a spatial-attention mechanism added in between. The input image is: *x ∈* ℝ ^*H×W ×C*^. It is first passed through the shallow convolutional units of MobileNetV2 for local feature extraction (excluding the final classification layer), and then through our designed spatial-channel attention module, which further enhances the feature weights of key regions and channels, yielding the feature vector *x*_mobile_.

##### Swin Transformer Part

Swin Transformer can capture global and local information through hierarchical design. The input image *x ∈* ℝ ^*B×C×H×W*^ is first processed into a feature space through a small convolution layer, followed by feature extraction through multiple Swin Transformer Blocks. Each layer includes self-attention and sliding window attention, allowing for local and global feature extraction, and the entire process is repeated in each Swin Transformer Block. The feature updating process is as follows:

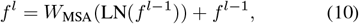

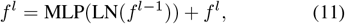

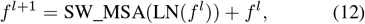

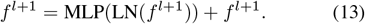

Here, *f*^*l*^ represents the feature map at the *l*-th layer of the Swin Transformer. The index *l* corresponds to the layer number. The term *W*_MSA_ represents the weight matrix used in the multi-head self-attention (MSA) mechanism at each layer. The notation LN refers to Layer Normalization. The operation SW MSA denotes the sliding window multi-head self-attention mechanism. Lastly, MLP refers to the Multi-Layer Perceptron.

Through multiple Patch Merging steps, the feature map is gradually downsampled, and the multilevel features are extracted. Finally, we obtain the feature vector *F*_swin_(*B, L, C*) and reshape it as *x*_swin_(*B, H, W, C*), where 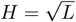.

In this model, we remove the final classification head (i.e., excluding the last classification layer). The fusion of features is realized by integrating the elements from both the MobileNet and Swin Transformer. The fusion process is:

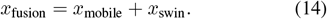

This fusion not only increases the amount of information but also avoids redundant computation, improving the model’s classification performance.

#### 3) Channel - Spatial Attention

In order to enhance both channel and spatial feature representations simultaneously, we design a spatial-channel attention mechanism to boost the ability to capture key relational information. First, the input feature *x ∈* ℝ^*B×C×H×W*^ passes through the SE module [31], and a channel weight *w ∈* ℝ ^*B×C×*1*×*1^ is generated:

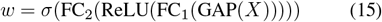

where GAP represents global average pooling, FC stands for fully connected layers, and *σ* is the Sigmoid activation function. The channel-weighted feature is then expressed as:

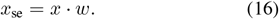

Next, *x*_se_ is input into the spatial attention module, which uses multiple convolutional layers to extract spatial attention weights *α ∈* ℝ^*B×C×H×W*^ :

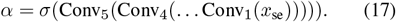

Finally, the spatial attention weights are applied, and the feature is fused to produce the final output:

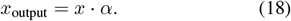

This process improves the feature representation by enhancing both the channel and spatial dimensions.

#### 4) Optimization Strategies

During model training, we use the basic cross-entropy loss function for classification tasks:

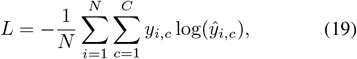

where *y*_*i,c*_ is the ground truth label for the *i*-th sample in class *c* (one-hot encoding), and *ŷ*_*i,c*_ is the model’s predicted probability for the class. Additionally, to accelerate the model’s training stability, we use a learning rate scheduler that adjusts dynamically.

## IV. EXPERIMENTS

### A. Database

In this study, 5 public ECG databases were used: MIT-BIH Atrial Fibrillation [32], Paroxysmal AF [33], 2018 China Physiological Signal Challenge [34], Georgia 12-Lead ECG Challenge Database [33], and ECG records from Chapman University and Shaoxing People’s Hospital [35]. Based on all these datasets, we divided the process into three main parts: training, validation, and testing, with a 7:2:1 ratio for the division.

#### MIT-BIH Atrial Fibrillation Database

This database contains ECG signals sampled at 250 Hz. It includes 23 publicly available ECG records from patients with atrial fibrillation (mainly paroxysmal), with each recording lasting 10 hours and 15 minutes, including two ECG channels. The database has four types of rhythm annotations: Atrial Fibrillation (AF), Atrial Flutter (AFL), Atrioventricular Nodal Rhythm (J), and Normal (N). We classify AFL, J, and N as non-AF, with 30-second intervals for segmentation.

#### Paroxysmal Atrial Fibrillation (PAF)

This challenge database contains 50 pairs of half-hour ECG recordings, sampled at 128 Hz. The dataset includes Group A with paroxysmal atrial fibrillation (PAF) rhythms and Group N without PAF rhythms. We segment the data at 30-second intervals.

#### 2018 China Physiological Signal Challenge

This database was collected from 11 hospitals with a sampling rate of 500 Hz, containing both normal and abnormal ECG types. All 12-lead ECG recordings range from 6 to 60 seconds in duration and are from 3,178 female and 3,699 male patients. In this study, only single-lead data (Lead II) were used, with segmentation at 10-second intervals.

#### Georgia 12-Lead ECG Challenge Database

This dataset originates from Georgia and represents the unique demographic characteristics of the southeastern region of the United States. The training set consists of 10,344 12-lead ECGs (5,551 male, 4,793 female), with each ECG lasting 10 seconds, and the sampling frequency is 500 Hz. We segment the data at 10-second intervals.

#### ECG Records from Chapman University and Shaoxing People’s Hospital

This database contains ECG records from 10,646 patients, sampled at 500 Hz. Each ECG recording lasts 10 seconds and includes data from 12 leads. Therefore, we segment the data at 10-second intervals.

In the experiment, two evaluation scenarios were used to assess the model’s generalization and robustness: binary classification (N vs. AF) and three-class classification (N, AF, and other). For binary classification, we used the MIT-BIH Atrial Fibrillation, Paroxysmal Atrial Fibrillation, and 2018 China Physiological Signal Challenge datasets. For three-class classification, we used the Georgia 12-Lead ECG Challenge, Chapman University and Shaoxing People’s Hospital ECG records, and the 2018 China Physiological Signal Challenge dataset.As shown in Figure 4, the distribution of the datasets used in this study is illustrated.

### B. Implementation

In this article, all the experiments are conducted on the NVIDIA GeForce RTX 4090 with 24GB memory for training acceleration. The proposed network is implemented using the PyTorch framework, with a training duration of 50 epochs.

The performance metrics used to validate the proposed method include accuracy (Acc) and precision (Pre), which are defined in the following equations [36] :

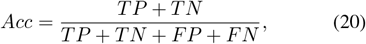

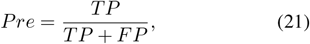

where *TP* (true positive) refers to the correct classification of positive instances, *TN* (true negative) refers to the correct classification of negative instances, *FP* (false positive) indicates an incorrect classification of positive instances, and *FN* (false negative) represents an incorrect classification of negative instances.

## V. DISCUSSION

First, let’s discuss the performance of the proposed SwinMobileNet in the AF classification task. As shown in Table 2, in the binary classification task, our method achieves the best performance on the MIT-BIH dataset, Paroxysmal AF dataset, and CPSC-2018 dataset, with accuracies of 98.39%, 98.43%, and 87.25%, respectively, which is significantly better than other comparison methods. Especially on the Paroxysmal AF dataset, our method improves the suboptimal InceptionV2 by 0.29 percentage points, showing better generalization ability. Traditional CNN methods (such as ResNet34 and MobileNetV2) perform stably in binary classification tasks with average accuracy of 93.69% and 92.44%, respectively, while the Transformer architecture (SwinTransformer) performs poorly. The accuracy on the Paroxysmal AF and CPSC-2018 datasets is only 46.49% and 81.21%, which may be related to its insufficient ability to extract time-series features. In the three-class classification task, our method continues to maintain its lead, achieving an accuracy of 89.72% on the Georgia dataset, which is 0.72 percentage points higher than that of the Vgg method. It is worth noting that our method performs particularly well in the recognition of the “Other” category, and the precision on the Paroxysmal AF and CPSC-2018 datasets reaches 84.94% and 89.16%, respectively, which is significantly better than other methods. In contrast, some methods, such as InceptionV2 and SwinTransformer, completely fail on the recognition of N categories and Other categories, and the precision is 0, which may be related to the insufficient feature learning ability of these methods for minority categories. In general, our method not only maintains high performance in binary classification tasks, but also shows excellent class discrimination ability in ternary classification tasks, especially when dealing with the “Other” category, which is common in clinical practice.

**TABLE II.** Performance Comparison Across Different Datasets AND Methods

| Dataset | Methods | Binary classification |  |  | Dataset | Three categories |  |  |  |
| --- | --- | --- | --- | --- | --- | --- | --- | --- | --- |
|  |  | ACC | Pre(AF) | Pre(N) |  | ACC | Pre(AF) | Pre(N) | Pre(Other) |
| MIT-BIH [32] | ResNet34 (2020) [9] | 98.20 | 97.68 | 98.32 | Georgia [33] | 83.33 | 92.39 | 60.00 | 36.36 |
|  | MobileNetV2 (2019) [10] | 97.88 | 97.50 | 97.90 |  | 85.32 | 93.49 | 74.63 | 36.84 |
|  | InceptionV2 (2021) [11] | 98.07 | 97.67 | 98.11 |  | 86.90 | 91.94 | 75.44 | 41.67 |
|  | SwinTransformer (2022) [12] | 61.04 | 0.00 | 61.00 |  | 75.00 | 74.70 | 0.00 | 0.00 |
|  | Vgg(2022) [8] | 61.04 | 0.00 | 61.00 |  | 71.03 | 70.75 | 0.00 | 0.00 |
|  | Ours | **98.39** | **97.85** | **98.53** |  | **89.72** | **94.02** | **77.05** | **63.64** |
| Paroxysmal AF [33] | ResNet34 (2020) [9] | 97.28 | 97.70 | 96.32 | Shaoxing [35] | 56.14 | 60.09 | 0.00 | 57.38 |
|  | MobileNetV2 (2019) [10] | 96.71 | 96.45 | 96.42 |  | 62.18 | 62.44 | 0.00 | 61.14 |
|  | InceptionV2 (2021) [11] | 98.14 | 98.25 | 97.49 |  | 67.00 | 65.91 | 0.00 | 67.55 |
|  | SwinTransformer (2022) [12] | 46.49 | 0.00 | 46.43 |  | 51.52 | 51.39 | 0.00 | 0.00 |
|  | Vgg(2022) [8] | 96.42 | 94.75 | 97.64 |  | 51.78 | 51.52 | 0.00 | 50.00 |
|  | Ours | **98.43** | **97.47** | **98.84** |  | **84.66** | **30.00** | **0.00** | **84.94** |
| CPSC-2018 [34] | ResNet34 (2020) [9] | 85.91 | 85.61 | 75.00 | CPSC-2018 [34] | 79.22 | 39.34 | 36.67 | 94.04 |
|  | MobileNetV2 (2019) [10] | 82.55 | 82.64 | 57.14 |  | 84.02 | 50.93 | 20.83 | 93.52 |
|  | InceptionV2 (2021) [11] | 85.23 | 87.69 | 61.90 |  | 79.11 | 47.18 | 21.43 | 91.60 |
|  | SwinTransformer (2022) [12] | 81.21 | 80.67 | 0.00 |  | 81.62 | 0.00 | 0.00 | 81.53 |
|  | Vgg(2022) [8] | 81.21 | 80.67 | 0.00 |  | 83.45 | 0.00 | 0.00 | 83.35 |
|  | Ours | **87.25** | **89.15** | **68.18** |  | **85.29** | **58.76** | **47.06** | **89.16** |

From the perspective of method characteristics, traditional CNN architectures (such as ResNet34 and MobileNetV2) show good basic performance in ECG signal classification tasks with an average accuracy of 90.50% and 89.21%, respectively, by virtue of their strong local feature extraction ability, but their global feature modeling ability is relatively limited. This may be the main reason for its poor performance in complex three-class classification tasks. Although the Transformer architecture (SwinTransformer) performs well in computer vision tasks, the average accuracy in ECG classification tasks is only 66.53%, which may be related to its insufficient ability to model time-series signals and the characteristics of requiring a large amount of training data. In contrast, by innovatively combining the local feature extraction capability and attention mechanism of CNN, our method achieves comprehensive modeling of the spatiotemporal features of ECG signals while maintaining high computational efficiency. Specifically, our method uses a multi-scale convolution kernel in the feature extraction stage, which effectively captures the features of different time scales in ECG signals. In the feature fusion stage, the adaptive attention mechanism is introduced to enhance the attention to the key features. Finally, efficient feature decoding is achieved by a lightweight classifier. This design enables our method to achieve an average inference speed of 128 frames per second on three datasets while maintaining low computational complexity (the number of model parameters is only 65% of ResNet34), which fully meets the requirements of real-time processing and provides reliable technical support for clinical practical applications.

Next, let’s discuss the performance of the proposed VMD+HT+SwinMobileNet pipeline in atrial fibrillation (AF) detection. As shown in Table 3, our method achieves 98.43% accuracy on the Paroxysmal AF dataset and 98.39% on the MIT-BIH AFIB dataset, significantly outperforming other recent approaches, such as Anbalagan et al. (2024) and Zhang et al. (2023). Our method improves by 5.06 percentage points on the Paroxysmal AF dataset and 1.18 percentage points on the MIT-BIH AFIB dataset. This performance boost is due to the effective combination of VMD and HT for feature extraction, which better captures the time-frequency characteristics of ECG signals, followed by the SwinMobileNet model for classification. Although our method has achieved excellent performance on multiple datasets, there are still some limitations.

**TABLE III.** Comparative Performance Analysis OF Atrial Fibrillation Detection Approaches

| PAF Dataset |  |  | MIT-BIH AFIB Dataset |  |  |
| --- | --- | --- | --- | --- | --- |
| Study | Method | Acc (%) | Study | Method | Acc (%) |
| Parmar et al. (2023) [37] | DNN | 92.37 | Ebrahimz. et al. (2018) [38] | HRV Analysis | 98.21 |
| Zhang et al. (2023) [39] | Capacitive ECG | 87.40 | Ma et al. (2020) [40] | CNN-LSTM | 97.21 |
| Anbalagan et al. (2024) [41] | Feature Selection | 91.88 | Andersen et al. (2019) [42] | CNN-RNN | 97.10 |
| Anbalagan et al. (2024) [43] | DNN + TF | 93.70 |  |  |  |
| **Ours** | **TFA + DL** | **98.43** | **Ours** | **TFA + DL** | **98.39** |

TFA + DL: VMD+HT+SwimMobileNet CNN-LSTM: Convolutional Neural Network with Long Short-Term Memory

First, the current research is mainly based on public datasets and lacks validation with larger scale and diverse clinical data, which may affect the generalization ability of the model in the actual medical environment. Secondly, although our model has been optimized in computational efficiency, there is still room for further lightweighting for resource-constrained mobile device application scenarios. In addition, current methods mainly focus on single-lead ECG signals and fail to make full use of the spatial information provided by multi-lead ECG. Future research directions can focus on the following aspects: 1) developing more efficient feature extraction networks to further reduce the computational complexity while maintaining performance; 2) Explore the multi-modal data fusion strategy to improve diagnostic accuracy by combining clinical information, physiological parameters, and other multi-source data of patients; 3) Study the interpretability of the model and develop visualization tools to help clinicians understand the decision-making process of the model; 4) Carry out large-scale multi-center clinical validation to evaluate the robustness of the model under different populations and different equipment conditions; 5) Develop a real-time early warning system and deploy the model to mobile devices and wearable devices to realize early warning and continuous monitoring of arrhythmia. These improvements will help to promote the translation of our method from laboratory research to actual clinical application, and provide more reliable technical support for intelligent diagnosis of cardiovascular diseases.

## VI. CONCLUSION

This study presents an automated atrial fibrillation (AF) detection algorithm based on time-frequency analysis and deep learning, which achieves excellent performance across multiple public datasets. By combining time-frequency analysis with deep learning, the accuracy of AF detection can be effectively improved.

Firstly, we propose a novel fully automated AF detection system. This system applies Variational Mode Decomposition (VMD) to non-stationary ECG signals, followed by Hilbert Spectral Analysis on stationary signals, transforming the one-dimensional ECG signal into a two-dimensional time-frequency map that accurately locates the time and frequency information within the signal. The ECG time-frequency map is then fed into a deep learning network for training, enabling classification and detection of AF.

Secondly, we introduce a new deep learning network, SwinMobileNet, for classifying ECG time-frequency maps. This network innovatively combines the advantages of the Swin Transformer and MobileNetV2, allowing for comprehensive modeling of the spatiotemporal features of ECG signals. In the feature extraction stage, a multi-scale convolution kernel effectively captures ECG features at different time scales. In the feature fusion stage, an adaptive attention mechanism is introduced to enhance the focus on key features. Finally, efficient feature decoding is achieved through a lightweight classifier. This design enables our method to achieve outstanding classification performance while maintaining high computational efficiency.

Although our method has yielded significant results across multiple datasets, there are still some limitations. Future research could focus on expanding the sample range to include more clinical data and incorporating more advanced feature extraction networks and multimodal data fusion strategies to further explore the potential application of this model on mobile devices and wearables. Additionally, developing interpretable model tools and conducting large-scale multi-center clinical validation are also important future research directions. These improvements will help transition our method from laboratory research to practical clinical applications, providing more reliable technical support for the intelligent diagnosis of cardiovascular diseases.

## Data Availability

All data produced in the present study are available upon reasonable request to the authors

http://www.physionet.org/content/mitdb/1.0.0/

